# Conceptualization of Health Literacy from a Nursing Perspective

**DOI:** 10.1101/2022.05.04.22274689

**Authors:** Angga Wilandika, Moses Glorino Rumambo Pandin

**Affiliations:** Faculty of Nursing, Universitas Airlangga, Surabaya, Indonesia; Faculty of Humanities, Universitas Airlangga, Surabaya, Indonesia; Faculty of Health Sciences, Universitas Aisyiyah Bandung, Indonesia

**Keywords:** health literacy, health education, nursing perspective, conceptualization

## Abstract

Health as an inseparable part of human beings needs to be maintained to achieve a complete human health degree. The role of health literacy in attaining optimum health is significant. When associated with nursing, health literacy must be interpreted as a part of the role and function of nursing. However, to understand health literacy, it is necessary to study it from the aspect of scientific formation itself and a nursing perspective. This review proposes an alternative conceptualization of health literacy from a nursing perspective. This review used an integrative search through four databases: ScienceDirect, ProQuest, SAGE Journal, and Google Scholar. Search using various combinations of keywords with the help of Boolean operators, including: health literacy, nursing perspective, nursing, and conceptualization combined as MESH terms. The inclusion criteria are peer-reviewed articles in English that discuss health literacy and nursing perspective. Articles published within the last six years (2017-2022). Research such as literature reviews, dissertations, editorials, commentaries, and other expert opinions are excluded. Ten articles were considered in this literature review. We describe the conceptualization of health literacy from the nurse’s point of view, the predictors that influence it, the dimensions surrounding health literacy, the implication of health literacy, and how nurses will participate in supporting this health literacy. In the end, this conceptualization will be used as an illustration material to integrate the concept of health literacy into various problems that become nursing tasks.

## INTRODUCTION

Health literacy is one of the important skills for nurses and patients in ensuring the quality of care that produces the best health outcomes. Health literacy is a concept that has developed for a long time, although in professional nursing practice, this concept is relatively new. In the scope of nursing so far, health literacy is interpreted in different ways, namely in health promotion.

Health promotion is one of the goals of nursing science. This activity is intended to maintain or improve individuals, groups, and communities (H. S. Kim, 2015). As stated by the American Nurse Association (ANA) (2015), nursing participates in ensuring the health of every individual, group, community, and society in general. Nursing as a profession must be able to protect, promote, and optimize the health and ability of each individual to achieve optimal health.

Health promotion in improving health literacy is one of the scopes of nursing work. Health optimization to improve health literacy must be understood as a form of embodiment of professional nursing practice. As one the health professionals that play a role in raising the issue of health literacy, nurses are very important to understand the forms of health literacy and the extent to which they can utilize this concept to support quality nursing care for patients.

Thus, to understand the nature of health literacy in nursing, how is the process of understanding health literacy. What are the goals or values contained in the concept of health literacy? It must be explained to provide a perspective on how nurses will use this concept. This literature review proposes an alternative conceptualization of health literacy from a nursing perspective. This review aims to describe the conceptualization of health literacy from the nursing point of view, the determinant factor that influences it, the dimensions surrounding health literacy, the implication of health literacy, and how nurses will participate in supporting this health literacy.

## METHODS

The study method used an integrative literature review. This review proposes an alternative conceptualization of health literacy from a nursing perspective. We systematically searched ScienceDirect, ProQuest, SAGE Journal, and Google Scholar. Search using various combinations of keywords with the help of Boolean operators, including: “health literacy” AND “nursing” AND “nursing perspective” AND “conceptualization”, combined as MESH term and keyword. The inclusion criteria applied in this study are peer-reviewed articles in English that discuss health literacy and nursing perspective. Articles published within the last six years (2017-2022). Research studies are carried out in various areas that specifically examine health literacy. Research such as literature reviews, dissertations, editorials, commentaries, and other expert opinions are excluded.

The first author performs an initial database search and articles for review. We used the PRISMA Flowchart 2009 (Moher et al., 2010) to record the article review and inclusion process (see Figure 1). An initial search of four databases yielded 423 results. After that, we collected all articles and removed duplicated articles. The source was excluded by title and abstract if it was not a peer-reviewed research study or related to health literacy in nursing scope. The next step was to narrow the selection of articles based on the year of publication and the research contexts. After the remaining articles were assessed for significant health literacy findings, 10 papers were selected for final inclusion (see Table 1).

**Table 1.** Characteristics of reviewed studies

| Author, Year | Research Methodology | Summary of findings |
| --- | --- | --- |
| Coşkun & Bebiş (2019) | D: quasi-experimental<br>S: 133 nursing students<br>V: health literacy, health-promoting lifestyle<br>I: Health-Promoting Lifestyle Profile (HPLP) and e-Health Literacy Scale (e-HEALS)<br>A: t-tests | Health literacy is an important aspect of achieving positive health behaviours. Health promotion learning can increase health literacy and positive health-related lifestyle behaviours. |
| Kim & Oh (2021) | D: cross-sectional<br>S: 558 nursing students<br>V: e-health literacy, health-promoting behaviours<br>I: e-Health Literacy Scale (heals) dan Health-Promoting Lifestyle Profile (HPLP-II) scale<br>A: a serial multiple mediation analysis | Electronic health literacy has a significant relationship with health promotion behaviour in students. The higher the health literacy, the better the health promotion behaviour. Health literacy does not directly increase health promotion behaviour but first encourages the use of social media in seeking information and then increases information-seeking behaviour and self-care agency. |
| Mosley & Taylor (2017) | D: cross-sectional<br>S: 15 lecturers, 53 nursing students<br>V: health literacy<br>I: health literacy scale<br>A: descriptive and qualitative analysis | Nurses must have the ability to adapt nursing care to patients with low health literacy. Health literacy education in the nursing education process is important to provide. |
| Kim et al. (2020) | D: randomized clinical trial<br>S: 250 pasien diabetes melitus tipe 2<br>V: health literasi knowledge, functional health literacy<br>I: Rapid Estimate of Adult Literacy in Medicine Scale, Test of Functional Health Literacy in Adults (TOFHLA) numeracy subscale<br>A: t-test, Chi-square test | Health literacy indirectly affects health outcomes. Self-efficacy and self-care skills are mediators between health literacy and the ability to control blood glucose levels and quality of life. |
| Wittenberg et al. (2018a) | D: cross-sectional<br>S: 74 nurses<br>V: communication, health literacy support<br>I: communication and health literacy support scale<br>A: descriptive and qualitative analysis | Most nurses reported communication challenges with patients who spoke English as a second language. Skills in providing health literacy support to patients are one of the core skills of nursing. |
| Ayaz-Alkaya & Terzi (2019) | D: cross-sectional<br>S: 303 nursing students<br>V: health literacy<br>I: Adult Health Literacy Scale<br>A: independent t-test and ANOVA | Nurses must have knowledge and experience in health literacy. The concept of health literacy should integrate into the nursing curriculum. Every nurse educator must emphasize the importance of health literacy and patient empowerment. |
| Nesari et al. (2019) | D: cross-sectional<br>S: 190 nurses<br>V: health literacy knowledge<br>I: Health Literacy Knowledge Experience Survey (HL-KES)<br>A: multiple linear regression | Nurses in this study were found not to have adequate knowledge and experience regarding health literacy practices. Efforts to increase equity in health literacy are very important in supporting the quality of patient care. |
| Turan et al. (2021) | D: cross-sectional<br>S: 232 nursing students<br>V: health literacy, healthy lifestyle behaviour<br>I: e-Health Literacy Scale in Adolescent, the Adolescent Lifestyle Profile Scale<br>A: Pearson correlation test, t-test, one-way ANOVA | Electronic health literacy is an important parameter in promoting healthy lifestyle behaviour in students. Health literacy positively affects interpersonal relationships, stress management, positive life perspective, spiritual health, and lifestyle. |
| Koduah et al. (2021) | D: cross-sectional<br>S: 876 nurses<br>V: health literacy knowledge<br>I: Health Literacy Knowledge and Experience Survey Instrument (HLKES)<br>A: ordinal logistic regression | Factors of personal characteristics, social and cultural values, nursing education, and experience have a relationship or connection with health literacy knowledge in nursing professional practice. |
| Munangatire et al. (2022) | D: Cross-sectional<br>S: 205 nursing students<br>V: health literacy<br>I: Understanding of Health Literacy (UHL) questionnaire<br>A: Pearson correlation test, t-test, one-way ANOVA | Nursing students understand health literacy concepts well, but all students must make more efforts to translate this understanding into health literacy skills. Health literacy skills can transfer to students' professional careers as nurses. |

Notes: D = design study; S = subject; V = variables; I = instruments; A = analysis

**Figure 1.**
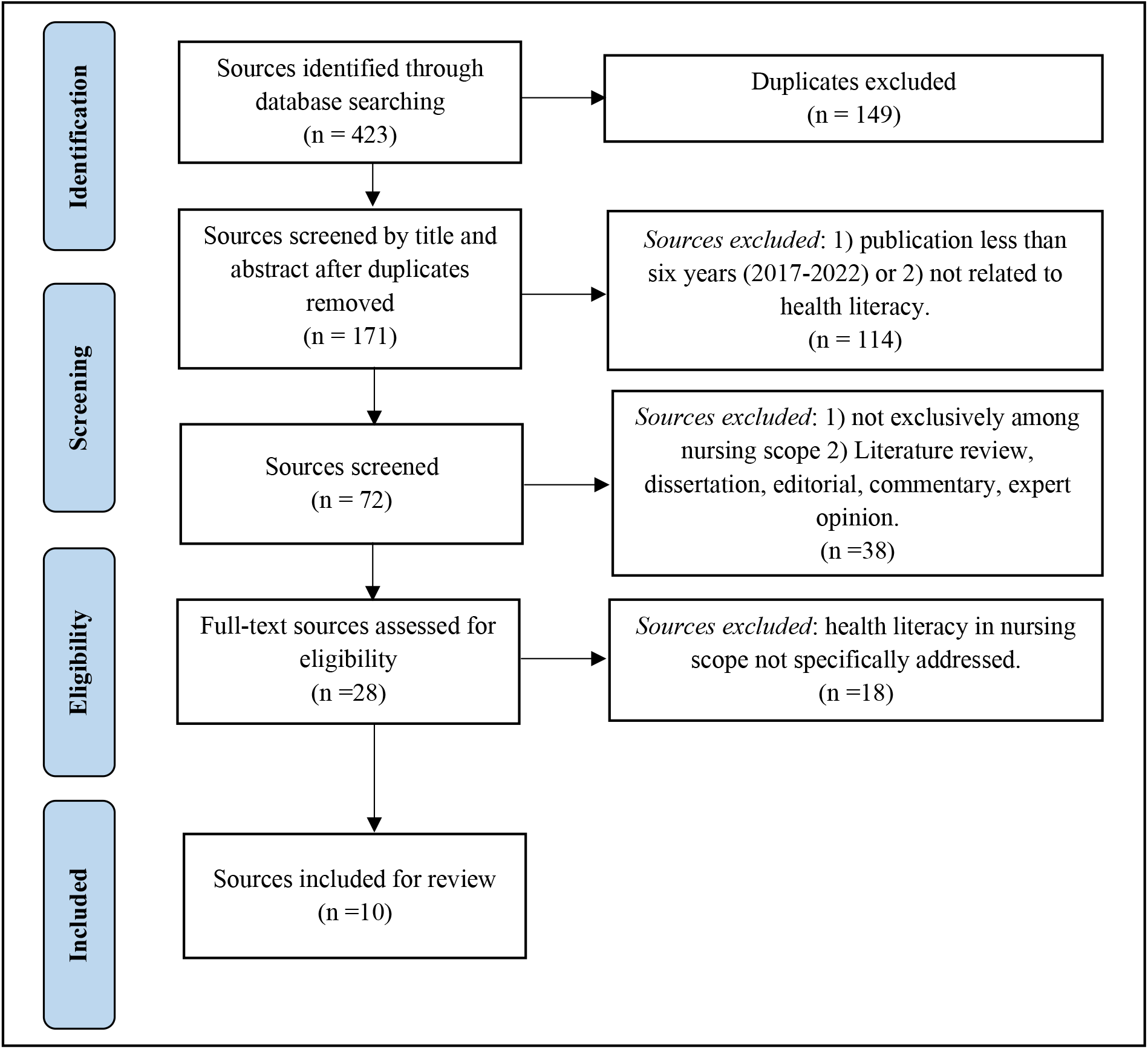
PRISMA Flow Diagram of article and inclusion

## RESULTS

The selection found 10 articles that had met the requirements, and further studies were carried out. The findings and conclusions of this study can be seen in Table 1. All articles reviewed discussed research on health literacy within nursing with research subjects including nurses, patients, and nursing students. Each article has information about health literacy used as the basis for research and the dimensions of health literacy used for research.

Health literacy is an important factor in the scope of professional nursing practice. Four studies found that health literacy has a role in achieving health behaviours and health promotion that impact health status (Coşkun & Bebiş, 2019; S. Kim & Oh, 2021; Koduah et al., 2021; Turan et al., 2021). Several studies have found that health literacy has an impact on improving healthy lifestyle behaviours and quality of life (Coşkun & Bebiş, 2019; M. T. Kim et al., 2020; Turan et al., 2021) and improving the quality of care while patients are hospitalized(Nesari et al., 2019). In patients, it also found that increasing health literacy impacted changes in better health status, such as changes in blood glucose levels in patients with diabetes mellitus (M. T. Kim et al., 2020).

Health literacy has a significant relationship with health behaviour. However, two studies reveal the impact of health literacy on health behaviour through intermediary mediating factors such as information seeking motivation, information-seeking behaviour, self-care agency, self-efficacy, and self-care management (M. T. Kim et al., 2020; S. Kim & Oh, 2021). One study revealed that health literacy does not directly increase health promotion behaviour but directly impacts the use of social media in seeking information, thereby increasing information-seeking behaviour and self-care agency (S. Kim & Oh, 2021). Meanwhile, Kim et al. (2020) said that self-efficacy factors and self-care management mediate between health literacy and health behaviour outcomes.

Health literacy within the scope of nursing care practice has a broad impact, limited to physical aspects and psychological, social, and spiritual aspects. Two studies reveal that health literacy skills impact the ability to cope with problems such as stress management, positive life thinking, and spiritual well-being (Coşkun & Bebiş, 2019; Turan et al., 2021). Thus, improving health literacy for nurses, nursing students, and especially patients becomes an important thing to improve to impact the development of the quality of nursing professional practice and health outcomes.

Four studies reveal that health literacy in nursing practice is one of the core skills of nurses (Coşkun & Bebiş, 2019; Munangatire et al., 2022; Nesari et al., 2019; Wittenberg et al., 2018a). Therefore, in nursing education process must be given health literacy knowledge to students (Ayaz-Alkaya & Terzi, 2019; Mosley & Taylor, 2017). Every nurse educator must emphasize the importance of health literacy skills for themselves and their patients. When students have graduated and work as nurses, they can apply them in a professional nursing career (Ayaz-Alkaya & Terzi, 2019; Munangatire et al., 2022).

In addition, each article reviewed in this study adopted the concept of health literacy, including dimensions, determinants, and consequences of health literacy, as can be seen in Table 2. Health literacy is a complex concept studied for many years, so it has various dimensions and is adapted to the context of specific health literacy. This study found that health literacy consists of traditional literacy, health literacy, information literacy, scientific literacy, media literacy, and computer literacy (Coşkun & Bebiş, 2019; S. Kim & Oh, 2021).

**Table 2.** Dimensions, factor determinants, and consequences of health literacy used in reviewed studies

| Reference | Dimensions | Factor Determinant | Consequences |
| --- | --- | --- | --- |
| Coşkun & Bebiş (2019) | • traditional literacy<br>• health literacy<br>• information literacy<br>• scientific literacy, media literacy<br>• computer literacy | Gender, BMI, family structure, parental knowledge status, income, smoking habit, alcohol drinking habit, history of chronic disease, drug use, and perception of health status | Improvement of healthy lifestyle behaviour. Improved ability regarding sources and how to find information related to health on the internet. They have an increased ability to determine which information is of good quality and which is not. |
| Kim & Oh (2021) | • information literacy<br>• scientific literacy<br>• media literacy<br>• computer literacy. | Age, gender, education, religion, duration of internet use, and health status | Health literacy increases the ability to use social media, online health information-seeking behaviour, self-care agency, and health behaviour. |
| Mosley & Taylor (2017) | • fundamental literacy<br>• scientific literacy<br>• civil literacy<br>• cultural literacy | Age, language, language ability and risk groups | Health literacy improves the patient's ability to interpret the disease process, thereby increasing the ability to manage their disease. Health literacy increases the capacity of individuals to make decisions regarding public health issues. Health literacy can increase the ability to recognize and understand the patient's cultural background to communicate effectively. |
| Kim et al. (2020) | health literacy knowledge<br>functional health literacy | Age, ethnicity, education, gender, disease severity | Health literacy interventions positively affect self-care skills such as knowledge, self-efficacy, compliance, and health status. Health literacy increases an individual's overall capacity to manage disease conditions effectively. |
| Wittenberg et al. (2018a) | health literacy support<br>perceived comport health literacy | Age, ethnicity, education, income, language, job position, demographic situation, work experience | Health literacy impacts patient health support and the quality of patient health. Nurses must have good communication skills, especially in assessing the level of health literacy; this is a strategy to support health literacy. |
| Ayaz-Alkaya & Terzi (2019) | individual's health information and medication usage<br>the status of knowledge about places of organs in the body | Age, gender, family income, social insurance, domicile, history of chronic disease, drug consumption, visual and hearing impairment, and internet use. | Nurses must have optimal health literacy skills to deal with rapid changes in the health care system. The higher the level of health literacy during undergraduate education, the more competent the nursing staff will be. Health literacy has an impact on increasing patient empowerment. |
| Nesari et al. (2019) | basic fact<br>health literacy screening<br>evaluation of written health care information<br>guidelines for written information<br>consequences of limited health literacy | Age, work experience, gender, and scope of the work area | Health literacy practices facilitate effective communication between health care professionals and the public. Health literacy practices increase public health literacy-related behaviours such as understanding health information and applying the information to navigate within the health care system and make informed decisions. |
| Turan et al. (2021) | individuals' knowledge<br>comfort and perceived skills at finding evaluating<br>applying electronic health information to health problems | Age, health insurance, expenses, health status, and internet usage. | Health literacy courses and healthy lifestyle behaviours are essential to support patients and families in accessing and using health information to improve patient safety and quality of care. In addition, nurses can use e-Health literacy and healthy lifestyle behaviours to improve the quality of care by supporting teaching methods and nursing role models. |
| Koduah et al. (2021) | basic facts<br>health literacy screening | Demographic factors: age, gender, marital status, and location of residence. Socio-cultural factors: religiosity, | Nursing practice must integrate health literacy knowledge professionally by taking characteristics, social and cultural values, nursing |
|  | consequences associated with low health literacy<br>guidelines written and health care materials<br>evaluation of health literacy intervention | cultural values, trust in others, group membership, income, and socioeconomic status. Educational and professional practice factors: nursing specialization, work experience, employment status, length of study, and higher education. | education, and experience into a personal account. |
| Munangatire et al. (2022) | access to health information<br>understanding health information<br>evaluating health information<br>utilizing health information | Age, marital status, gender, and medical history. | When they become professional nurses, efforts should be made to ensure that students understand the importance of health literacy and health practice for themselves and their patients. |

The two studies above have similarities in literacy dimensions with the study conducted by Mosley & Taylor (2017). Studies on health literacy apply four dimensions of health literacy: fundamental literacy, scientific literacy, civic literacy, and cultural literacy. Meanwhile, health literacy can also be seen from the knowledge and functional aspects of health, such as the literacy dimension in Kim et al. (2020) research, which includes health literacy knowledge and functional health literacy. The same thing is seen in Wittenberg et al. (2018a) study and Turan et al. (2021). Ayaz-alkaya & Terzi (2019) apply several dimensions of health literacy, namely health literacy support, perceived comfortable health literacy, individuals’ knowledge, comfort, and perceived skills at finding, evaluating and applying electronic health information to health problems.

Meanwhile, two other studies that refer to the dimensions of health literacy are based on the basic concepts of literacy which are specifically related to the research context. The dimensions of health literacy include basic health literacy facts, health literacy screening, evaluation of the effectiveness of written health care information, guidelines for written health care information, and consequences of limitations (Koduah et al., 2021; Nesari et al., 2019). In contrast to the study of Munangatire et al. (2022), his research applies the dimensions of health literacy, namely access to health information, understanding health information, evaluating health information, and utilizing health information.

Health literacy in nursing practice is also influenced by various determinant factors, as shown in Table 2. The determinant factors found to affect health literacy include age, gender, education, language, income, and marital status (Ayaz-Alkaya & Terzi, 2019; Coşkun & Bebiş, 2019; M. T. Kim et al., 2020; S. Kim & Oh, 2021; Koduah et al., 2021; Mosley & Taylor, 2017; Munangatire et al., 2022; Wittenberg et al., 2018a). In addition, increasing patient health literacy is influenced by factors using information media, duration of internet use (Ayaz-Alkaya & Terzi, 2019; S. Kim & Oh, 2021; Turan et al., 2021), disease status, disease history, and disease severity (Ayaz-Alkaya & Terzi, 2019; Coşkun & Bebiş, 2019; M. T. Kim et al., 2020; S. Kim & Oh, 2021; Munangatire et al., 2022; Turan et al., 2021). In addition, ethnic, socio-cultural, religiosity, and experience factors in a nurse’s job affect health literacy (M. T. Kim et al., 2020; Koduah et al., 2021; Nesari et al., 2019).

## DISCUSSION

### Health Literacy in Nursing Perspective

Health literacy is a process that requires knowledge, motivation, and competence to access, understand, assess, and apply health information. This process is carried out to access, understand, assess, and use health information that enables a person to improve health quality and prevent the emergence of disease (Coşkun & Bebiş, 2019). Sørensen et al. (2012) also said that decision-making in health care, disease prevention, and health promotion is done by accessing, understanding, assessing, and using appropriate health information. According to Ancker et al. (2020), the definition of health literacy is the extent to which individuals can obtain, process, and understand basic health information needed to make informed decisions.

Meanwhile, the Healthy People 2030 framework states that health literacy is the extent to which a person can find, understand, and use this information to make decisions about the health of themselves and others based on that information (US Department of Health & Human Services, 2020). Someone who has good health literacy will be able to participate in public and private dialogue about health, medicine, scientific knowledge, and cultural beliefs (Zarcadoolas et al., 2005). Based on the explanation above, health literacy emphasizes the ability of people to use information, not only to understand it.

In the research of Mosley and Taylor (2017), health literacy is an ability that nurses must possess in adapting nursing care to patients. Health literacy is the extent to which individuals can use the information studied appropriately to determine the most appropriate nursing intervention given to patients. Nurses are also expected to assess the level of health literacy of patients (Wittenberg et al., 2018a). In addition, decisions made based on health literacy are highlighted findings because being “well informed” is not just the right decision. Thus, health literacy is a person’s process of obtaining and using that information based on appropriate information in making decisions regarding the health of oneself and others.

Health literacy can apply at the individual and community levels in public health. Further links between health literacy and public health, McKenna et al. (2017) conceptualize that health literacy is a relational concept that involves how individuals interact with health and social systems. This concept is so complex that it requires a greater understanding of the context of the public health experience. The same thing was expressed by Guzys et al. (2015) defines health literacy from the perspective of public health and should emphasize the knowledge and skills needed to prevent disease and improve health in everyday life.

The definition of health literacy is evolving towards a focus on public health promotion and empowerment that allows everyone to have control over their health. Health literacy also helps identify an individual’s motivation and ability to access, understand, and use the information to promote and maintain health (Nutbeam et al., 2018). Health literacy is an essential aspect of health promotion, including environmental, political, and social aspects that can hinder or improve health outcomes (World Health Organization Regional Office for Europe, 2019). Health literacy at the community level can increase knowledge, skills, and health behaviours that result in better health impacts (Guzys et al., 2015). So, health literacy is a form of an individual, group, or community’s ability to obtain information, understand, evaluate, and apply it to overcome health-related problems. In the end, they can make health-related decisions based on the correct information.

### A Determinant Factor of Health Literacy

Within the scope of nursing, health literacy skills as the basis for professional nursing practice in supporting nursing care for patients are influenced by various determinant factors. Interventions to improve health status through a good health literacy that occurs in individuals, communities, and society must consider the determinants of health literacy. According to Koduah et al. (2021), determinant factors that affect health literacy consist of demographic factors (age, gender, marital status, location of residence), socio-cultural factors (religiosity, cultural values, trust in others, group membership, income, social-economic status), and educational and professional practice factors (occupational specialization, work experience, employment status, length of study, educational attainment).

This determinant is in common with Sørensen et al. (2012), which said the predictors for health literacy are divided into personal, social-environmental, and situational factors. Individual factors include age, gender, race, socioeconomic status, education, work status, occupation, income, and knowledge. Following social and environmental factors have demographic conditions, culture, language, political power, and social systems as for the situational factors that affect health literacy, namely social support, family and groups, the use of media, and the physical environment.

Demographic or personal factors, especially age, gender, income, education, and employment status, were found to have a close relationship to determining a person’s health literacy (Ayaz-Alkaya & Terzi, 2019; Coşkun & Bebiş, 2019; M. T. Kim et al., 2020; S. Kim & Oh, 2021; Koduah et al., 2021; Munangatire et al., 2022; Wittenberg et al., 2018a). Among these individual factors, age has the closest relationship with health literacy. The higher the age, the lower the health literacy (Do et al., 2020; Liu et al., 2015; Yang et al., 2021). Likewise, Sántha’s (2021) study found that health literacy significantly correlated with age and education factors. However, in his research, these two factors are categorized into socioeconomic factors. In addition, Yang et al. (2021) said that in addition to personal factors, access to information or the use of information sources such as the internet is a factor that determines a person’s literacy. Health literacy is a concept based on the existence of the information, so access to information becomes an inseparable part of the formation of health literacy. Literacy skills will not be sufficient for someone with less access to information. This statement is supported by several studies that found that using various sources or access to information will determine a person’s level of health literacy (Ayaz-Alkaya & Terzi, 2019; S. Kim & Oh, 2021; Turan et al., 2021). Motivational factors and the individual’s ability to access information also affect health literacy (Izadirad & Zareban, 2022).

Meanwhile, Guo et al. (2021) found that sociodemographic factors (gender, education, income, and access to information) were not directly related to health literacy. Social-democratic relationships were observed through personal factors such as self-efficacy, social support, and health status. All of these individual factors have a positive relationship with health literacy. Health status in sick people is a close predictor of health literacy. Health status, disease history, disease severity, other health disorders, and the use of drugs will fundamentally affect a person’s efforts to increase or decrease health literacy (Ayaz-Alkaya & Terzi, 2019; Coşkun & Bebiş, 2019; M. T. Kim et al., 2020; S. Kim & Oh, 2021; Munangatire et al., 2022; Turan et al., 2021).

In addition to the factors mentioned above, spiritual and environmental factors also affect health literacy. Ecological aspects of the residence, education, place of work, and religious beliefs impact health literacy (Pronk et al., 2021). The study conducted by Koduah et al. (2021) found that religiosity factors affect health literacy. Nurses who have firm religious beliefs view that having health is important in their lives, so having adequate health literacy becomes an inseparable part. It is the same with the home environment (Ayaz-Alkaya & Terzi, 2019; Koduah et al., 2021; Wittenberg et al., 2018a) and the work environment (Koduah et al., 2021; Nesari et al., 2019) as nurses also determines individual health literacy and actions to increase patient health literacy. These factors will determine the health literacy of individuals, groups, or communities.

### Dimensions of health literacy

Health literacy contributes to a person’s health status. Health literacy puts the health of oneself, family, and society into the context of knowledge, understanding the factors that influence it, knowing how to get it, and knowing how to use it. A person with adequate health literacy will be competent in paying attention to their health, families, and communities (Zhang et al., 2021).

From the perspective of public health, health literacy generally accommodates the concept of health through three health domains, namely health services, disease prevention, and health promotion. As expressed by Sørensen et al. (2012) regarding the conceptual model of health literacy that health literacy is a multidimensional concept and consists of different components. The health literacy model also identifies individual and system-level factors influencing a person’s literacy level. In addition, in the health literacy concept model, a pathway connects health literacy with health outcomes. The competencies needed to achieve health literacy goals include access, understanding, appraising, and applying information.

The four dimensions of health literacy mentioned above are the same as those used by Munangatire et al. (2022) in their research, which consists of access to health information, understanding health information, evaluating health information, and utilizing health information. These dimensions are used to assess health literacy in the scope of health practice in nursing.

1. Access

Access refers to seeking, finding, and obtaining information in health literacy. The ability to acquire and access knowledge depends on understanding, time, and trust. At this stage, the ability to access information must balance in measuring the accuracy of the sources of information found (Matterne et al., 2021). If the source of information accessed and obtained is inadequate or has an inappropriate tendency, it will harm understanding and making decisions at the health literacy application stage. Thus, determining the source of information is also an important factor in accessing information.

2. Understand

Understanding competence in health literacy means the ability to understand accessible health information. Understanding information depends on expectations, perceived utility, individualization of results, and interpretation of causality (Sørensen et al., 2012). Understanding of information can also be influenced by emotions, such as when people face a problem related to a deteriorating health condition or disclosure of an illness (McKenna et al., 2017).

3. Appraise

Assessment is the ability to interpret, filter, assess, and evaluate health information that has been accessed. The processing and assessment of health literacy are influenced by the complexity of the information, jargon, and partial understanding of the information. Dehghani (2021) states that the evaluation of health information is critical in applying health literacy. If the review of health information is not carried out properly, it will cause difficulties in using the information as a basis for decision-making. However, the ability to assess the information obtained is still determined by the previous stage, namely the ability to read information comprehensively.

4. Apply

Application is the competence or ability to communicate and use the information to make decisions in improving or maintaining health status. The application of health literacy to obtain effective communication depends on a thorough understanding. Information that is successfully received, understood, and evaluated regarding the truth of the information, when applied, will produce the best decision because of good news.

The health literacy process produces knowledge and skills that enable a person to achieve health goals (Matterne et al., 2021). Health literacy is a form of knowledge, competence, and motivation to access, understand, assess, and apply health information in making the right decisions in health care, disease prevention, and health promotion. However, health literacy, a complex concept that has been widely developed in various studies, has various perspectives related to its dimensions.

Several studies use the dimensions of health literacy developed by Zarcadoolas et al. (2005), namely fundamental literacy, science literacy, civic literacy, and cultural literacy (Mosley & Taylor, 2017), and the development of traditional literacy, health literacy, information literacy, scientific literacy, media literacy, and computer literacy (Coşkun & Bebiş, 2019; S. Kim & Oh, 2021) as a basis for assessing health literacy according to the research context. Fundamental literacy refers to skills and strategies that involve the ability to read, converse, write, and count (Zarcadoolas et al., 2005). Fundamental literacy is also the main aspect that affects daily life and impacts patients’ ability to interpret health information and education (Mosley & Taylor, 2017).

The second dimension is science literacy, which is fundamental knowledge of scientific concepts, understanding of technology, and technical complexity, awareness of scientific uncertainty, and acceptance of changes in science (Zarcadoolas et al., 2005). According to Mosley and Taylor (2017), low scientific literacy can cause patients to misinterpret the disease process, which has the potential to reduce their disease management ability. Understanding basic health information is necessary for managing health problems, although some medical conditions require a higher level of health literacy.

Civic literacies, which is the third dimension according to Zarcadoolas et al. (2005), are media literacy skills and awareness that individual decisions can impact public health. Meanwhile, the last dimension, namely cultural literacy, relates to recognizing and using collective beliefs, habits, broad perspectives, and social identities to interpret and act on health information (Zarcadoolas et al., 2005). Everyone needs to recognize and understand the patient’s cultural background to communicate effectively. Based on cultural literacy, an understanding of how the professional culture of health care providers and patient culture will affect communication (Mosley & Taylor, 2017).

On the other hand, Wittenberg et al. (2018a), Turan et al. (2021), and Ayaz-Alkaya & Terzi (2019), in their research, apply the dimensions of health literacy based on the scope of the problem study. The context of study includes health literacy support, perceived comport health literacy (Wittenberg et al., 2018a), individuals’ knowledge, knowledge about places of an organ in the body (Ayaz-Alkaya & Terzi, 2019), comfort and perceived skills at finding evaluating, dan applying electronic health information to health problems (Turan et al., 2021). Meanwhile, the study of Nesari et al. (2019) and Koduah et al. (Koduah et al., 2021) apply the dimensions of health literacy as an indicator in assessing health literacy based on the instrument developed by Cormier (2006). The indicators include basic facts, health literacy screening, consequences of low health literacy, written guidelines, health care materials, and evaluation of health literacy interventions.

The various dimensions of health literacy that already exist are alternative solutions to solve a problem or develop scientific concepts related to health literacy. The multidimensionality of health literacy implies that everyone who will use the health literacy approach must conduct an in-depth study first in using the dimensions of health literacy. The development of health literacy in nursing requires assessment so that the concepts used are appropriate and able to answer every problem.

### Implications and role of nurses in health literacy

Health literacy has been widely recognized as a mediator between individual and social health status and health outcomes. Health literacy is the initial component of obtaining a permanent health culture and optimal health status (Barton et al., 2018). Health literacy allows a person to make a good decisions on health care, disease prevention and health promotion to maintain and improve health (Alsubaie & Salem, 2019; Guo et al., 2021). Adequate health literacy will improve healthy lifestyle behaviours and increase individual capacity to manage disease conditions (Coşkun & Bebiş, 2019; M. T. Kim et al., 2020; Mosley & Taylor, 2017).

Meanwhile, limited health literacy will affect people’s behaviour related to self-management of chronic conditions, outcomes of health efforts (Sántha, 2021), and health costs in society (Dehghani, 2021). Several studies have also revealed that inadequate health literacy can lead to poor health implications, unsuccessful preventive care, and high health care costs. Health literacy also affects the effects of social determinants of health and health status (Mantwill et al., 2015; Rasu et al., 2015). This condition shows that inadequate health literacy will lead to health disparities.

The health literacy gap has a massive impact on overall health disparities. Efforts to reduce health literacy gaps are essential to reduce health disparities, especially at the community or community level (Gómez et al., 2021; Pronk et al., 2021). Health literacy affects a person’s health attitudes and behaviour, as in the research by Silva & Santos (2021), which found that health literacy has a relationship with knowledge and attitudes toward preventing Covid-19. The better health literacy, the better the perspective on preventing disease Health literacy is not passive but a form of participatory ability and foundation in positive behaviour change. Individuals with an adequate level of health literacy can take responsibility for their health and the well-being of their families and communities (Pitts & Freeman, 2021). Therefore, efforts to strengthen health literacy can enhance primary health conditions at the individual and community levels. Increasing health literacy will also improve overall health and welfare status.

Low levels of health literacy are also associated with poor health (Balmer et al., 2020). The application of health literacy that is carried out at any time and in all patient care settings will impact achieving client empowerment, engagement, activation, and optimal health outcomes (Loan et al., 2018). Nurses have a significant role in educating patients about health information and health promotion (Mosley & Taylor, 2017). Providing health literacy support to patients is also a nurse’s core skill (Wittenberg et al., 2018b). In carrying out this role, nurses need to have a broad understanding of health literacy.

In nursing, health literacy is an essential mediator in communication strategies for all clients. Clients cannot fully participate in their health care or make appropriate health decisions without accurate information for their health literacy needs (Ballard & Hill, 2016). Nurses as health workers who have a closer relationship with clients can become facilitators to meet these health literacy needs through health education or health promotion activities.

A nurse who plays a role in improving health literacy must understand the extent of health literacy skills and the level of understanding that the patient has of a disease or problem. Sometimes nurses often overestimate the client’s level of health literacy, report using their feelings, and only rely on the client’s level of education as an indicator in concluding the client’s literacy ability (Dickens et al., 2013; Macabasco-O’Connell & Fry-Bowers, 2011; Parnell, 2014). That actions taken by the nurse became problematic.

Nurses cannot assume a person’s literacy level is only seen from external appearances such as age, education, or socioeconomic status. When the nurse provides information to the client, the nurse must be careful in interpreting the client’s body language movements, such as a nod of the head, lest this is interpreted as an indication that the patient has understood the information provided (Parnell, 2014). As previously explained, health literacy is not a simple concept of understanding but something that must assess carefully, persistently, and intensely. Health literacy that is studied inadequately will eventually become an obstacle to health literacy for clients. As stated by Cohen et al. (2013), in ensuring understanding and overcoming the problems of health literacy barriers, nurses must carry out adequate assessments to assess the client’s level of knowledge, emotional reactions to information, and involvement from outside or other health teams that can provide support.

Wittenberg et al. (2018b) provide several recommendations in assessing health literacy in individuals or groups, or communities who experience health literacy barriers, including 1) nurses must understand the beliefs and cultural norms of clients; 2) repetition of information explanations or providing information more technically adapted to the cultural approach of the client; 3) nurses must be able to observe verbal and nonverbal cues, and must be careful with medical terms that may be new to clients, or use simple language in providing information; and 4) someone who has a language barrier can involve a translator. Therefore, the ability of nurses to assess patient understanding and address challenges related to health literacy needs is vital to providing quality nursing care so literacy can improve.

## CONCLUSIONS

The findings show the conceptualization of health literacy from a nursing point of view. Health literacy is defined as a process and skill in using information resources to make health decisions, individually, in groups, and society. Determination of decisions related to health is based on the ability to find, understand, assess, and evaluate the information obtained.

Nurses must also understand that health literacy is a complex and multidimensional concept. Using a health literacy approach to overcome the problems faced must consider the concept’s dimensions, determinant factors, and implications. By fully understanding the concept of health literacy from a nursing point of view, nurses can use this approach to solve problems related to literacy and ultimately improve the quality of nursing care for patients.

## Data Availability

All data produced in the present work are contained in the manuscript

## ACKNOWLEDGEMENTS

We express our most profound appreciation to all those who have contributed to this study.

